# Using COVID-19 deaths as a surrogate to measure the progression of the pandemics

**DOI:** 10.1101/2020.09.27.20202564

**Authors:** Carlos Hernandez-Suarez, Efren Murillo-Zamora

## Abstract

The IFR (Infection Fatality Risk) is one of the most important parameters of an infectious disease. If properly estimated, the observed number of deaths divided by the IFR can be used to estimate the current number of infections and, if immunity is permanent, we can estimate the fraction of susceptible which can be used to plan reopening of activities and vaccine deployment, when these become available. Here we suggest how to use the observed deaths by COVID-19 in an arbitrary population as a surrogate for the progression of the epidemic with the purpose of decision making. We compare several estimates of IFR for SARS-CoV-2 with our estimate that uses the number of additional deaths in households in a database population of 159,150 laboratory-confirmed (RT-qPCR) COVID-19 by SARS-COV-2 in Mexico. The main result is that if the number of deaths in a region is close to 2 per thousand individuals, the fraction of remaining susceptible may be too small for the vaccine to make a difference in the total number of infected.

## 1. Introduction

COVID-19 pandemic has shown to have a low lethality, nevertheless, the burden of the disease so far is huge. Economical activities are suspended or reduced and there is pressure for reopening of schools which requires more information, not only available to policymakers but also to the general public, to ameliorate civic unrest. The competition for developing a vaccine is keen, with about 321 vaccine candidates with 32 in clinical trials in progress (Le et al., 2020), raising safety concerns (Harrison and Wu, 2020; Peeples, 2020), and the additional economic cost for the purchase and deployment of vaccines has not been yet added to the burden of the disease.

At the moment, without vaccines or effective pharmaceutical treatments available, the decision on whether reopening activities or not depends mostly on the number of additional infections/deaths that will be caused because of some policy, say, opening schools, allowing public gatherings, opening touristic places, increasing the current density allowed in some places as cinemas, restaurants, buses, and planes, etc. For a highly infectious virus as SARS-CoV-2, the decision must be rooted in the amount of remaining susceptible in the region affected for some change in policy.

Using the number of individuals that access private of public hospitals as a surrogate of the current number of infections, or even the number of confirmed cases, is not accurate, because those quantities strongly depend on the availability of medical services that are not always accessible to the bulk of a population in many countries, or simply because of lack of resources for test deployment or because of policies that disregard testing. The number of deaths is less dubious. if we manage to calculate the average number of infections that will result in the death of the average individual, then, we can use this number to estimate the number of infections that were required to observe the current number of deaths, and from here, the share of susceptible available in a well-defined population. Even although deaths were not reported as COVID-19 related, records may exist that shed light on the likelihood that death was or not caused by SARS-CoV-2 infection, which is part of the policy adopted by some countries like Belgium, which is, without doubt, one of the reasons why this country keeps one of the highest number of deaths per capita in Europe.

Before proceeding, we need to deploy two facts. The first one attempts to establish that, with few exceptions, it is almost impossible to stop the pandemic by mitigation and control measures, and the most we can do is reducing the infection rate (at a huge economic and social cost) which is known as *flattening the curve*. The second establishes that we are in the condition now to have reliable lower bound estimates for IFR, the Infection Fatality Risk of SARS-CoV-2. It is the confluence of these two facts that allow us to establish a surrogate for the progression of the epidemic in a region and thus, for the calculation of the share of susceptible available, which is the basis to decide on reopening activities and vaccine deployment.

### Fact 1: SARS-CoV-2 is highly contagious and will infect most of the population

Several studies have shown that the basic reproductive number *R*_0_ is high (Sanche et al., 2020; D’Arienzo and Coniglio, 2020; Najafimehr et al., 2020; Liu et al., 2020; Alimohamadi et al., 2020), and it has been calculated as high as 5.7. It is important to remember that *R*_0_ is the potential of disease transmission in the absence of any control or mitigation measures, and it is the potential that comes into play as soon as control or mitigation measures are suspended. We calculated the *R*_0_ for 40 countries using only the first two weeks of data, analyzing the progression in the number of deaths. These 40 countries were the first countries affected by pandemics except for China. The *R*_0_ calculated is shown in Figure 1. The estimation of the true potential of the disease is more evident for those countries that were affected first (e.g. Italy and Spain), where our estimate of *R*_0_ is about the same as the reported by Sanche et al. (2020). The learning curve is evident in the reduction of *R*_0_ when the pandemic advances. Estimates of *R*_0_ in countries where the pandemic arrived at later stages will not reflect the true potential of the virus alone, but the preparedness of a population.

**Figure 1:**
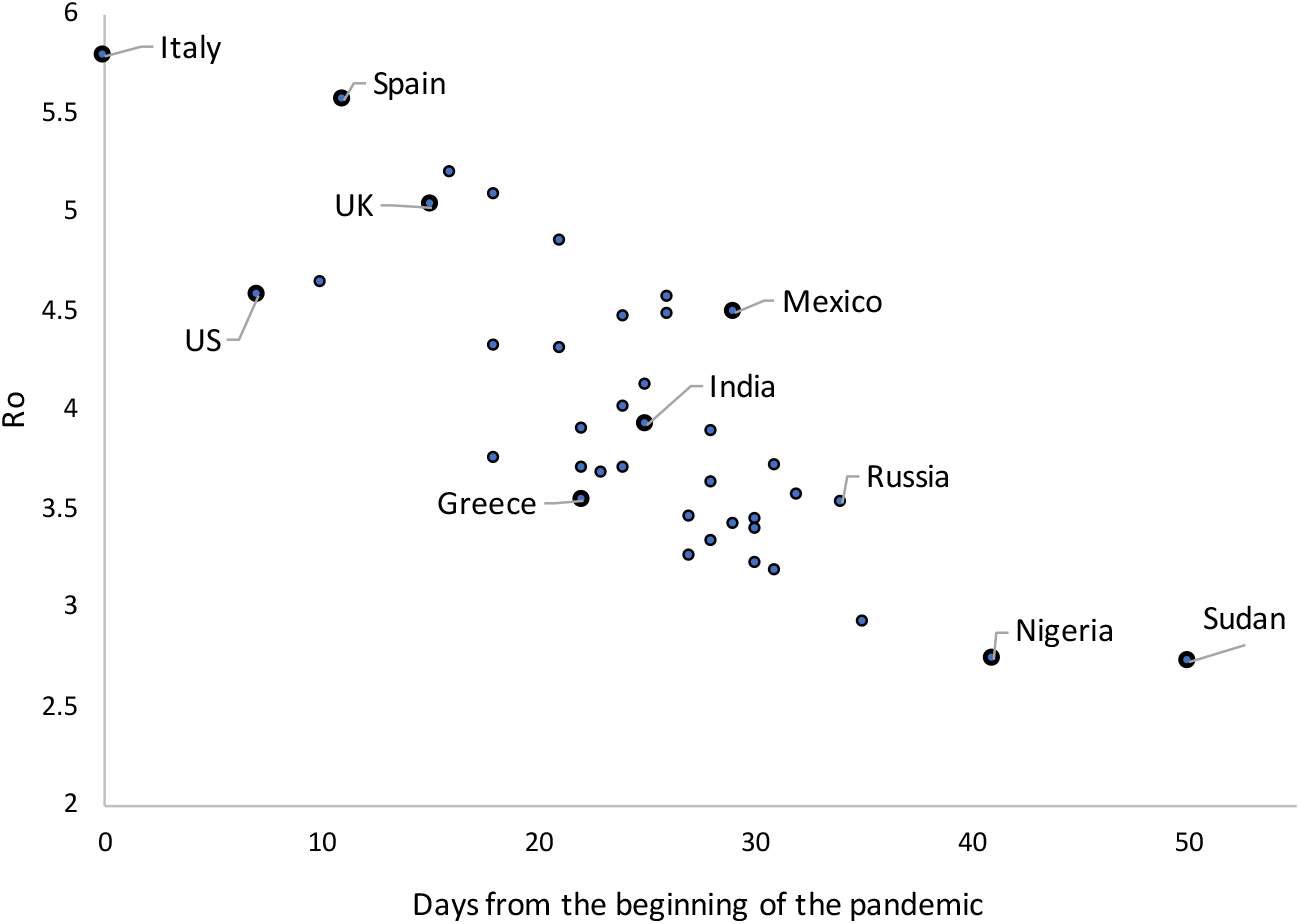
Age of the pandemic vs. *R*_0_. For each country, the baseline was taken as the date when the accumulated number of deaths was at least 5. Day zero is 02/24/2020. See Table 1 for expanded data.

Table (1) in Appendix contains the main statistics derived from the analysis of *λ* and *R*_0_ for the countries analyzed. From the estimated *λ*’s, we can see that the first countries to be affected had an *R*_0_ larger than 5 whereas this was decreasing to achieve values just above 2.5, this implies that at the beginning *λ* was as high as one effective contact every three days to one effective contact every 5 days, on average.

**Table 1:** Observed frequencies of deaths per household. The index case is excluded.

| Deaths | Households |
| --- | --- |
| 0 | 130,248 |
| 1 | 921 |
| 2 | 41 |
| 3 | 22 |
| 4 | 4 |
| 5 | 2 |
| 6 | 1 |
| 7 | 1 |

There are some facts we need to consider when a virus spreads with that strength: first, forecasting the size of the epidemics (total number of infected) is simpler, since social contact structures are bypassed and become irrelevant, and thus the contact network resembles more a random mixing pattern. For this random mixing, the estimated fraction of infected *f*, can be approximated (Kermack and McKendrick, 1927) with:

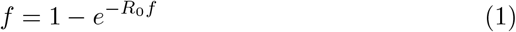

(for a probabilistic derivation, see Hernandez-Suarez and Mendoza-Cano, 2009). For instance, if *R*_0_ = 5.7 the fraction of infected is *f* = 0.99. However, if we manage to reduce *R*_0_ by half, *f* = 0.93, which is not a big difference in the size of the epidemic. Nevertheless, reducing *R*_0_ by half is an incredible task that involves mainly lockdowns and face-masks, and the former has a huge economic cost that can not hold for long except for rich countries or countries whose political or cultural organization allows its implementation. Nevertheless, even if a country manages to implement actions to reduce *R*_0_ to a value smaller to one, and can support citizens economically to maintain the lockdown for long periods, a handful of infected individuals that enter the country from abroad can restart the infection process, if lockdowns are lifted. From here, the observed waves (see Figure 2 for examples in Japan, Cuba, S. Korea and New Zealand). This is particularly true for a disease where the number of asymptomatic individuals is by far, larger than that of symptomatic and thus, difficult to control. As a consequence, equation (1) can be seen as a quota that must be fulfilled, as long as the fraction of remaining susceptible is larger than *f*.

**Figure 2:**
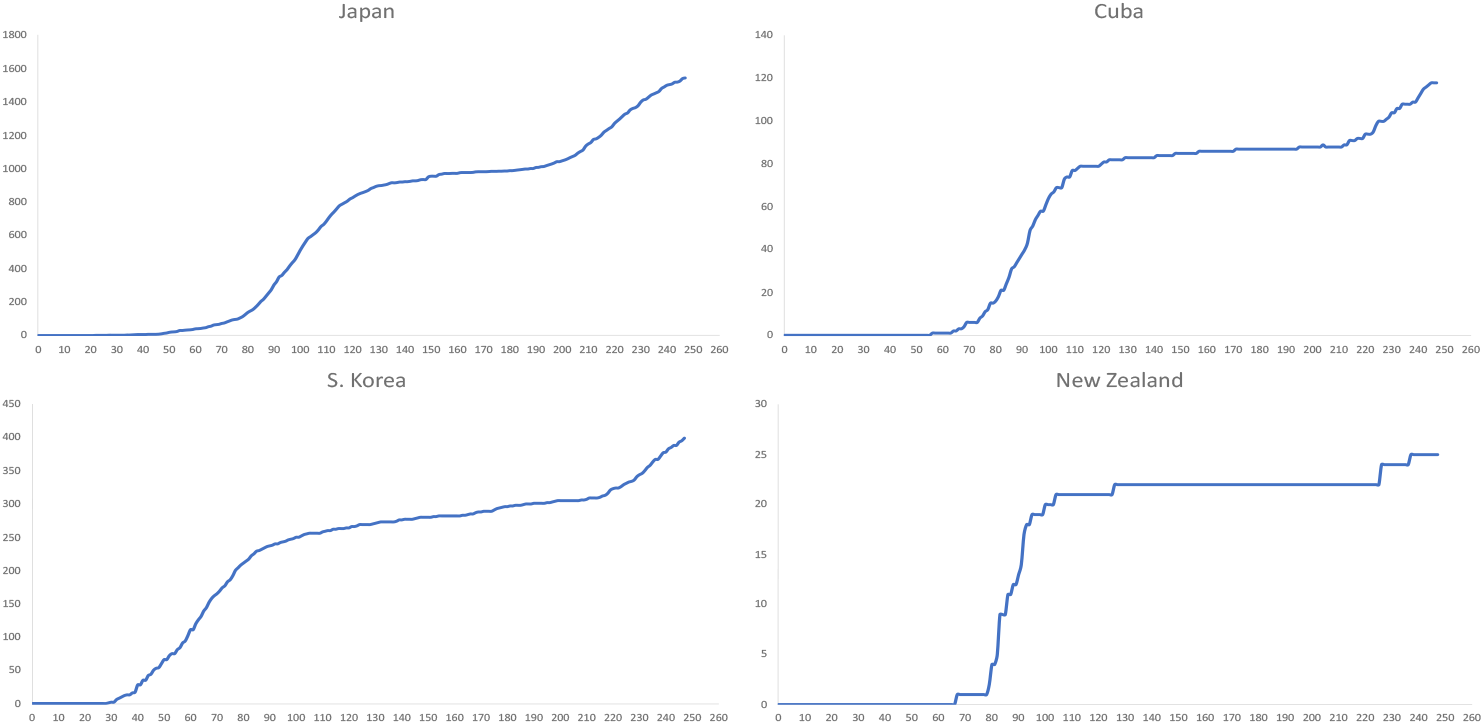
The appearance of a second wave. Cumulated deaths per day. The use of deaths instead of confirmed cases delays the detection of a wave but is more efficient in the sense that does not depend on testing policies, availability or panic. The current IFR^*\**^ (per million inhabitants) is Japan: 14; Cuba: 11; S. Korea: 9; New Zealand: 5. Data updated to 10/29/2020.

### Fact 2: the overall IFR of SARS-CoV-2 is somewhere between 0.2 and 0.3 %

All calculations in this section are done with the data on deaths reported to 10/29/2020 (Dong et al., 2020). Also, in this section we use IFR to indicate the true overall Infection Fatality Risk of SARS-CoV-2, that is, it is the fraction of individuals that die if infected with SARS-CoV-2, whereas IFR^*\**^ indicates some estimate of IFR for a region of interest. Many attempts have been made to estimate this important parameter that would be a remarkable surrogate to measure the progress of the epidemics in an arbitrarily defined region or population, but the main inconvenient is that while the number of deaths from SARS-CoV-2 can be approximated, the denominator, the total number of infected from the disease is elusive, especially considering that a large fraction of individuals that are infected are asymptomatic. Besides, the efficacy of immunity tests to detect who has been infected and recovered has been challenged with the findings that the ability to detect antibodies is reduced in a few days, especially in those with mild or no symptoms (Long et al., 2020; Ibarrondo et al.). Recently, Eyre et al. (2020) reported that a high fraction of individuals with none or mild symptoms may be undiagnosed mainly due to the calibration strategies of some standardized tests and concluded that samples from individuals with mild/asymptomatic infection should be included in SARS-CoV-2 immunoassay evaluations.

Some studies to estimate the IFR have been recently released for Iceland (Gudbjartsson et al.), India (Mukhopadhyay and Chakraborty, 2020), Germany (Streeck et al., 2020) and Denmark (Erikstrup et al., 2020) among others. In a review of 23 studies where the IFR was estimated Ioannidis (2020) a median of 0.25% is reported, which is about 2.5 deaths for each 1000 individuals. The study in Iceland, (Gudbjartsson et al.), the most comprehensive to date, proposes an IFR of 0.3% (95% CI, 0.2 to 0.6). Special care must be taken with this later estimate, since Iceland has a population of 321,641, with only 29 deaths so far. In other studies that used transmission models fed with serological data, the IFR estimates where as high as 2.5-5 (Thorne, 2020) and 3-8 (Roques et al., 2020) per thousand infections. Wilson (2020) estimated the IFR using serological data from New York City and arrived to an IFR estimate of 8.6. We believe this particular estimate is too high, and will support our claim analyzing the behavior of the epidemic in some regions. The reason of such a high estimate may be the sampling scheme, which was not random at all, and would explain the high discrepancy between the observed differences in the number of infected between males and females (15.9 and 12 percent respectively) which the authors acknowledge they can’t explain.

#### Estimation of the IFR using an alternative method

Hernandez-Suarez et al. (2020) proposed an alternative method to estimate the IFR based on the assumption that all those living with an infected individual will be infected. We use the same data set that has grown from 3,193 households to 131,240 households. The essence of the method is as follows: let *n*_*j*_ be the number of individuals living in a house where an infected was confirmed infected with SARS-CoV-2, whose symptoms started on day *d*_*j*_. Let *x*_*j*_ the number of deaths among the remaining *n*_*j*_ − 1 individuals whose symptoms started after day *d*_*j*_, then, an estimate of the IFR is:

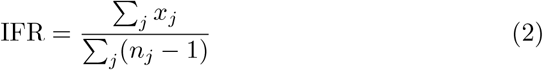

In this example we build an approximation to (2) using a database from Mexico’s IMSS (Instituto Mexicano de Seguro Social), the Mexican Institute for Social Insurance. The database has 212,935 (laboratory-confirmed (quantitative reverse transcription polymerase chain reaction, RT-qPCR) COVID-19 by SARS-COV-2 cases from April 1 to September 17, 2020. In an attempt to consider only households with final outcomes we excluded cases with symptoms onset in the last 6 weeks before the final case recorded, that is, we considered only cases with symptom onset from April 1 to August 6, 2020. The final data set has 135,855 cases. The mean age of this final set was 47.9 years with a standard deviation of 16.5 years and median 47 years. From these, there were 54 % males and 46 % females. Tables 2 and 3 contain summary statistics from the final data set.

**Table 2:** Observed frequencies of deaths by gender

| Age | Males | Females |
| --- | --- | --- |
| 0-9 | 0 | 0 |
| 10-19 | 546 | 578 |
| 20-29 | 7,956 | 8,324 |
| 30-39 | 13,413 | 12,818 |
| 40-49 | 13,770 | 12,399 |
| 50-59 | 12,728 | 9,688 |
| 60+ | 18,716 | 13,329 |
| Total | 67,129 | 57,136 |

**Table 3:** Summary statistics of data set

|  | N | % |
| --- | --- | --- |
| **Total** | 135,855 |  |
| **Gender** |  |  |
| Male | 73,256 | 53.9 |
| Female | 62,599 | 46.1 |
| **Condition** |  |  |
| Ambulatory | 75,725 | 55.7 |
| Hospitalized | 60,130 | 44.3 |
| **Outcome** |  |  |
| Recovered | 103,432 | 76.1 |
| Death | 32,423 | 23.9 |

Every individual has an associated NSS (Social Security Number) that is shared among all members of a family. As an approximation, we assumed all individuals sharing the same NSS live in the same household, and then grouped the cases in households. If there were more than one case in a household, we only considered the household if all cases were already solved a deaths or recoveries. In every household with more than one case we consider the index case as the individual with the earliest symptom onset and counted the number of deaths among the remaining members of the house. From the final set of 131,240 households, there were 130,248 with no deaths among the remaining members of the household and 992 houses with at least one additional death (see Table 1).

From Table 1 we have Σ_*j*_ *x*_*j*_ = 1, 108 We do not have Σ_*j*_ *n*_*j*_ and we build an estimate with the number of households times the average household size, that is, 131, 240 *× µ* where *µ* = 3.7 (Encuesta Intercensal, 2015). Using (2) we have our estimate of IFR is

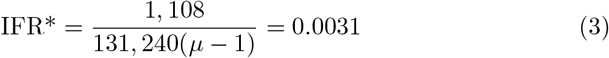

that is, 3.1 cases per thousand infected, which is close to 2.5, the median of 23 studies reported in Ioannidis (2020) and very similar to the 3.0 recently derived from the study in Iceland (Gudbjartsson et al.) Special care must be taken with our estimate (3) since we are building a rough approximation to the total number of additional individuals living in each household.

If the disease will affect most of the population, as suggested previously, then, the most effective way to estimate the overall IFR is by measuring the fraction of deaths in a population in which the epidemic has evolved for a long time and it is over or near the end. It is evident that we need to focus in regions with: *a*) high observed IFR^*\**^ and *b*) lack of new waves. Such is the case of regions as New Jersey and New York states (see figure 3). The current IFR^*\**^ calculated for these states is 1.854 and 1.729 respectively.

**Figure 3:**
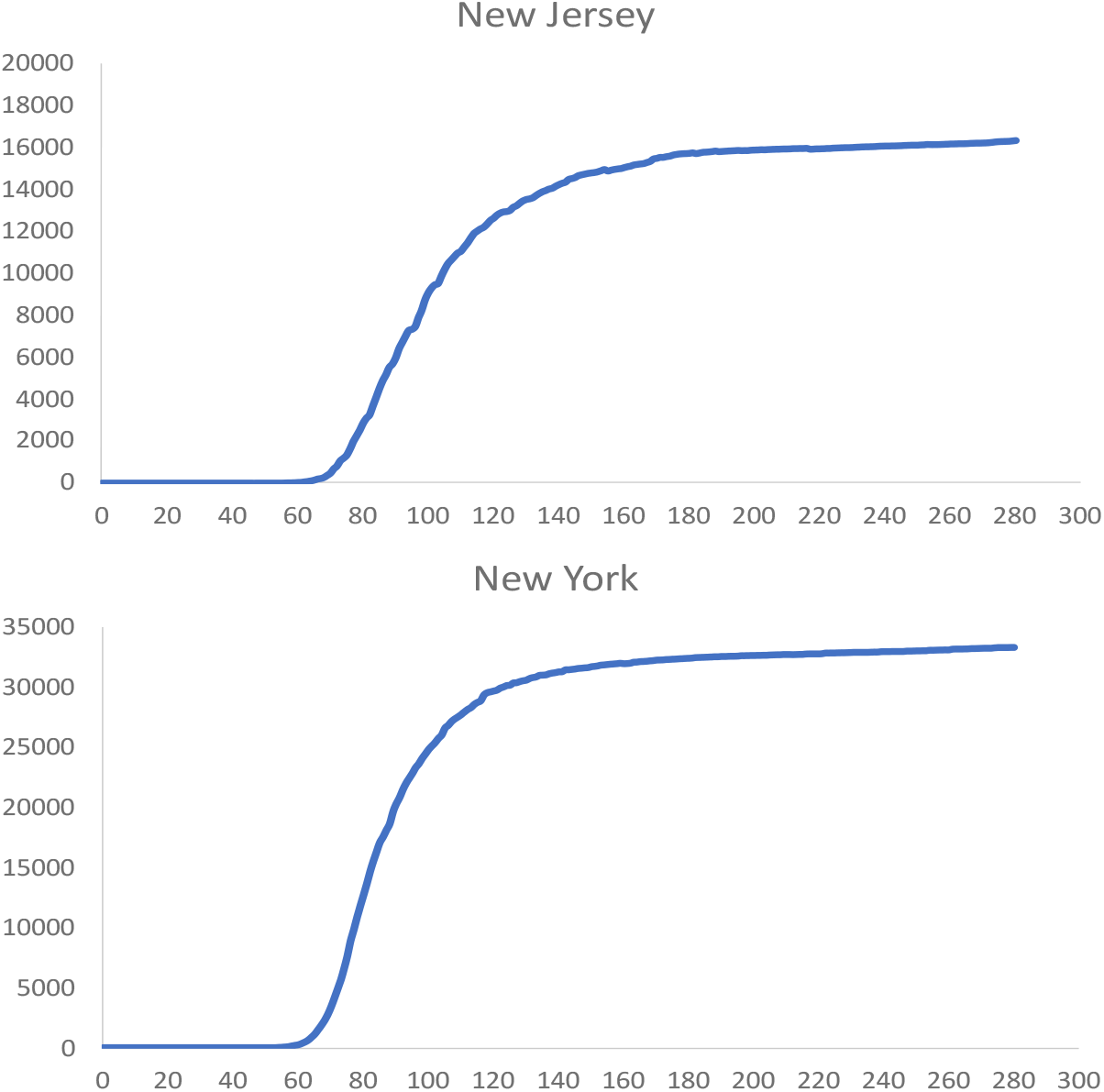
Absence of second waves in the states of New Jersey and New York. Cumulated deaths per day. The current IFR^*\**^ (per million inhabitants) is New Jersey: 1.854; New York: 1.729. Data updated to 10/29/2020.

The lack of new waves in these two states indicates that the virus has infected and killed a fraction of individuals close to the IFR, and thus the epidemic rampaged those states and it stopped due to what is commonly called *herd immunity*, that actually conveys no immunity at all, but a lack of a significative mass of susceptible for the *R*_0_ to be effective. Thus, the progression of the epidemic in a region can be estimated with *IFR*^*\**^/*IFR*.

## 2. Consequences from the previous facts for vaccines and the return to normality

As a consequence of the two previous facts, we can conclude that it is possible to monitor the development of the epidemic in a region by following the number of deaths. One can estimate the fraction of total infected at time *t*, asymptomatic or not, with:

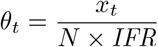

where *x*_*t*_ is the number of observed deaths at time *t* and *N* is the population size. The estimated fraction of susceptible as a function of the IFR in New Jersey and New York is shown in figure 4. We can see that if IFR=2/1000, the fraction of remaining susceptible in New Jersey and New York is 7.3% and 13.5% respectively, nevertheless, if it is as high as 3, the fraction of susceptible raises to 38% and 42% for those states.

**Figure 4:**
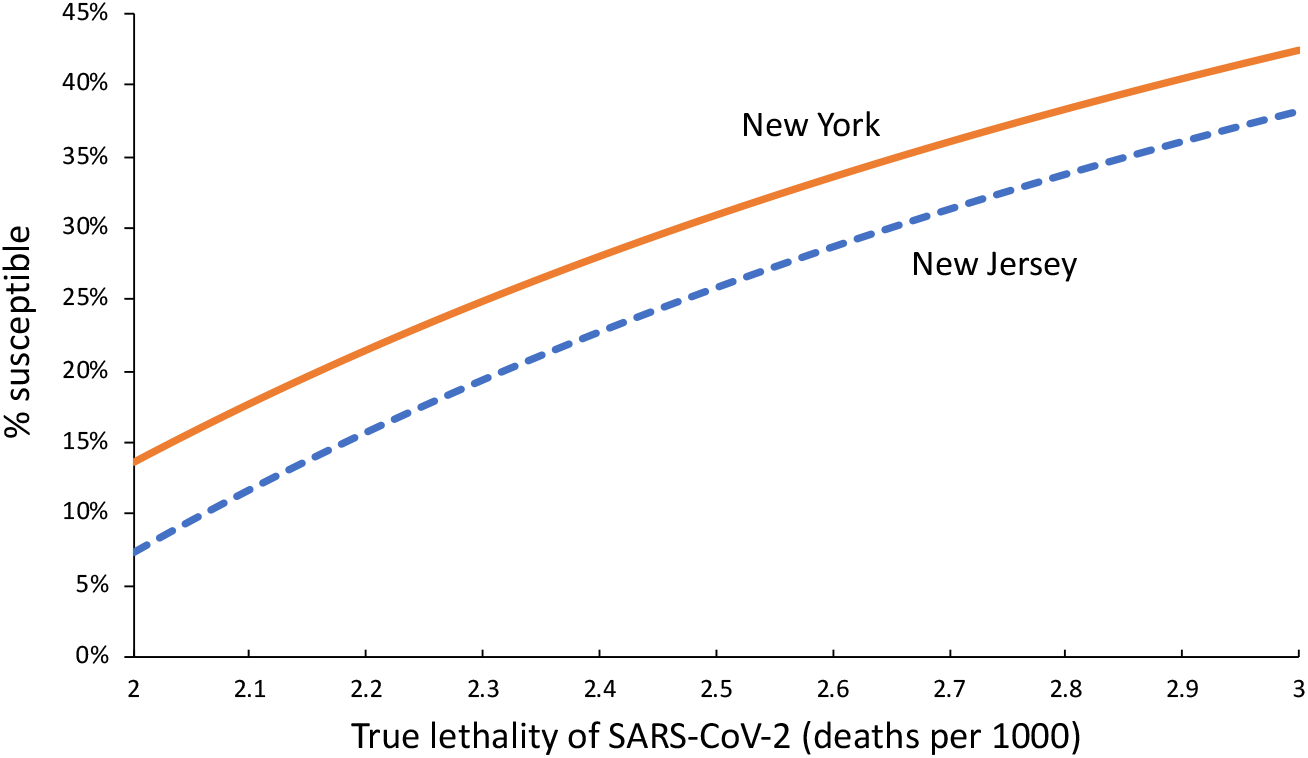
The fraction of remaining susceptible in the states of New Jersey and New York as a function of the lethality of SARS-CoV-2.

## 3. Discussion

Because of the aforementioned facts, it is very important to monitor the COVID-19 related deaths and every effort has to be made to improve current estimates of the IFR. Hernandez-Suarez et al. (2020) suggested that in light of the high contagiousness of SARS-CoV-2, the secondary cases resulting in fatalities in a household with at least one confirmed infected, may be useful to estimate the IFR, which would provide a large amount of data and minimal testing requirements. Antibody testing in New York shows to date (Data updated to 10/29/2020) that out of 2,608,997 tests, 642,977 were positive for antibodies (NYC-Health) which implies 24% of the population may be immune, contradicting our findings that for an IFR as high as 2.5 per thousand the percentage of infected should be close to 70% already (see Figure 4). Nevertheless, the fraction of people with antibodies is not a good surrogate of the fraction of infected with SARS-CoV-2 because it underestimates the number of infected for two reasons: first, antibody testing is voluntary and it is natural to expect that individuals with no symptoms are less compelled to be tested than those symptomatic, so, a large fraction of infected is not tested, and second, we already mentioned some failures in testing reported, apparently due to calibration procedures that tend to fail in individuals with none or mild symptoms (Eyre et al., 2020).

The idea that the share of susceptible in New Jersey is relatively small, is supported on the fact that the number of active cases reported to date (10/29/2020) is close to 42,000 and unless they are fully quarantined the infectious pressure from these individuals and those unreported must be huge, nevertheless, no new significant peak is observed. The same happens in New York, although the share of susceptible is higher.

Everything leads to the following question: what will be the purpose of a vaccine in a region where the IFR^*\**^ is already large enough to suggest there is only a small fraction of susceptible with limited chance of infection? This question is especially valid from the comprehensive study in Iceland that strongly suggests the existence of immunity due to infection (Gudbjartsson et al.). Unless it is proven that immunity wanes beyond a protective level after some time, everything seems to indicate that those regions where the share of estimated susceptible is already low, should have less priority in the distribution of vaccines. If facts 1 and 2 are not taken into account, a vaccination campaign in a region where the proportion of deaths among the population is close to the IFR, will just give the impression of being effective but will not play a role, regardless of the efficacy of the vaccine.

## Data Availability

No open data available

## Acknowledgements

This work is part of the DFID-UNHCR-World Bank program “Building the Evidence on Protracted Forced Displacement: A Multi-Stakeholder Partnership”. The program is funded by UK aid from the United Kingdom’s Department for International Development (DFID), it is managed by the World Bank Group (WBG) and was established in partnership with the United Nations High Commissioner for Refugees (UNHCR). The scope of the program is to expand the global knowledge on forced displacement by funding quality research and disseminating results for the use of practitioners and policy makers. This work does not necessarily reflect the views of DFID, the WBG or UNHCR.

## 5. Appendix

**Table A1:** Contact rate *λ* and *R*_0_ for the first 40 countries with diagnosed SARS-CoV-2. *R*_0_ was calculated at the beginning of the epidemic in each country.

| Country | $\lambda$ | $R_0$ | $R^2$ | Country | $\lambda$ | $R_0$ | $R^2$ |
| --- | --- | --- | --- | --- | --- | --- | --- |
| Italy | 0.366 | 5.79 | 0.997 | Philippines | 0.237 | 3.748 | 0.988 |
| Spain | 0.352 | 5.569 | 0.995 | Colombia | 0.235 | 3.708 | 0.996 |
| Netherlands | 0.329 | 5.196 | 0.998 | Denmark | 0.234 | 3.696 | 0.994 |
| Germany | 0.322 | 5.083 | 0.999 | Poland | 0.234 | 3.694 | 0.992 |
| United Kingdom | 0.319 | 5.036 | 0.995 | Norway | 0.233 | 3.681 | 0.997 |
| Belgium | 0.306 | 4.844 | 0.996 | Romania | 0.229 | 3.627 | 0.987 |
| France | 0.294 | 4.645 | 0.979 | Chile | 0.226 | 3.569 | 0.996 |
| US | 0.29 | 4.579 | 0.993 | Greece | 0.224 | 3.545 | 0.994 |
| Peru | 0.289 | 4.571 | 0.996 | Russia | 0.223 | 3.528 | 0.996 |
| Mexico | 0.284 | 4.494 | 0.998 | Pakistan | 0.218 | 3.45 | 0.993 |
| Turkey | 0.283 | 4.48 | 0.998 | Israel | 0.218 | 3.444 | 0.995 |
| Brazil | 0.282 | 4.461 | 0.997 | Morocco | 0.216 | 3.418 | 0.993 |
| Switzerland | 0.273 | 4.323 | 0.995 | Ukraine | 0.215 | 3.393 | 0.995 |
| Sweden | 0.273 | 4.31 | 0.982 | Panama | 0.211 | 3.333 | 0.991 |
| Ecuador | 0.26 | 4.116 | 0.997 | Hungary | 0.206 | 3.252 | 0.994 |
| Austria | 0.254 | 4.009 | 0.992 | Slovenia | 0.204 | 3.222 | 0.998 |
| India | 0.248 | 3.925 | 0.99 | Finland | 0.201 | 3.18 | 0.994 |
| Portugal | 0.248 | 3.922 | 0.992 | UA Emirates | 0.185 | 2.926 | 0.993 |
| Canada | 0.247 | 3.903 | 0.996 | Nigeria | 0.173 | 2.732 | 0.995 |
| Ireland | 0.246 | 3.888 | 0.992 | Sudan | 0.173 | 2.73 | 0.995 |

